# Mitochondrial DAMPs as mechanistic biomarkers of mucosal inflammation in Crohn’s disease: Study protocol for prospective longitudinal cohort study in Scotland

**DOI:** 10.1101/2022.03.21.22270313

**Authors:** Shaun Chuah, Rebecca Hall, Emma Ward, Broc Drury, Gwo-tzer Ho

## Abstract

The MUSIC study is a multi-centre, longitudinal study set in the real world IBD clinical setting to investigate and develop a new biomarker approach utilising mitochondrial DAMP and circulating DNA in assessing mucosal healing in Crohn’s disease. We present the study protocol (approved by East of Scotland Research Ethics Service, Scotland, United Kingdom - Reference No. 19/ES/0087) on 17^th^ September 2019 and registered in ClinicalTrials.gov as NCT04760964 for this on-going project based in Scotland, UK.

## INTRODUCTION

### 1.1 Background

Inflammatory bowel disease (IBD) comprising of Crohn’s disease (CD) and Ulcerative Colitis (UC) are common, chronic incurable immune-mediated conditions affecting the gastrointestinal tract. The incidence and prevalence of IBD are increasing, currently affecting 300 000 individuals in UK and 10 million worldwide. CD can affect anywhere along the gastrointestinal tract, mostly commonly the last part of the small bowel (terminal ileum) and the right side of the large bowel. CD is characterized by aphthous ulcers (appearances like common mouth ulcers). UC on the other hand, affects only the large bowel and has a diffuse, continuous inflamed appearance typified by easy bleeding from the gut lining. Both conditions are associated with debilitating symptoms and signs such as excessive tiredness, uncontrolled bowel habit, abdominal pain, weight loss, malnutrition. In severe cases, IBD patients can develop abdominal abscesses, bowel perforation and sepsis.

### 1.2 The current problem

IBD is a chronic progressive inflammatory condition. Despite recent progress in identifying factors that increase one’s likelihood to (e.g. genetics), or that can trigger (e.g. drugs, stress) a flare, we do not understand why gut inflammation seen in IBD does not resolve (in comparison to other forms of gut inflammation for example, infectious gastroenteritis). Because of uncontrolled gut inflammation, IBD patients are at risk of long term bowel wall damage and the serious complications associated with this. For example, in CD, 50% of patients will develop strictures (narrowing of the bowel), fistulas (abnormal connections between different parts of the bowel or to other organs) or abscesses (pockets of pus) that require surgery within 10 years of diagnosis.

Medical treatments have improved with more options available. Current clinical approach follows a sequential route of conventional medical treatments with corticosteroids, immunomodulators, tumour necrosis factor (TNF) inhibitors and the newer biologics in that order. However, there is significant inter-individual variation in therapeutic response and our current best standard approach involving azathioprine and TNF-inhibitors are effective in at best 50% of CD patients with severe CD^1^. Loss of anti-TNF response occurs in 20-30% of CD patients within one year of treatment initiation. We lack the knowledge (or tools) underlying such molecular heterogeneity in clinical presentation and treatment response in both UC and CD.

### 1.3 What is the MUSIC study?

The MUSIC study is a multi-centre, longitudinal study set in the real world IBD clinical setting to investigate and develop a new biomarker approach that aims to inform both patients and clinicians of the current state of the affected gut lining (how inflamed or whether the bowel wall has completely healed).

This new biomarker approach will study a panel of molecular signs in IBD patients’ blood, stools and biopsies that will be correlated to the current gold standard of direct gut visual examination using ileo-colonoscopy and flexible sigmoidoscopy tests (a fibre-optic examination of the lower small bowel and large bowel).^1^ Here, the state and appearances of IBD patients’ gut lining will be assessed over 1year in response to current standard drug treatment given to them by their NHS IBD consultant.

This approach will focus on the role of damage associated molecular patterns (DAMPs), also known as ‘danger signals’. DAMPs are found in our own cells and are released during tissue stress or injury. Like signals from bacteria, they can trigger inflammation. In the MUSIC study, we will use blood, stool, saliva and gut samples obtained from participants during active IBD and in clinical remission in order to understand how DAMPs contribute to the development of gut inflammation.

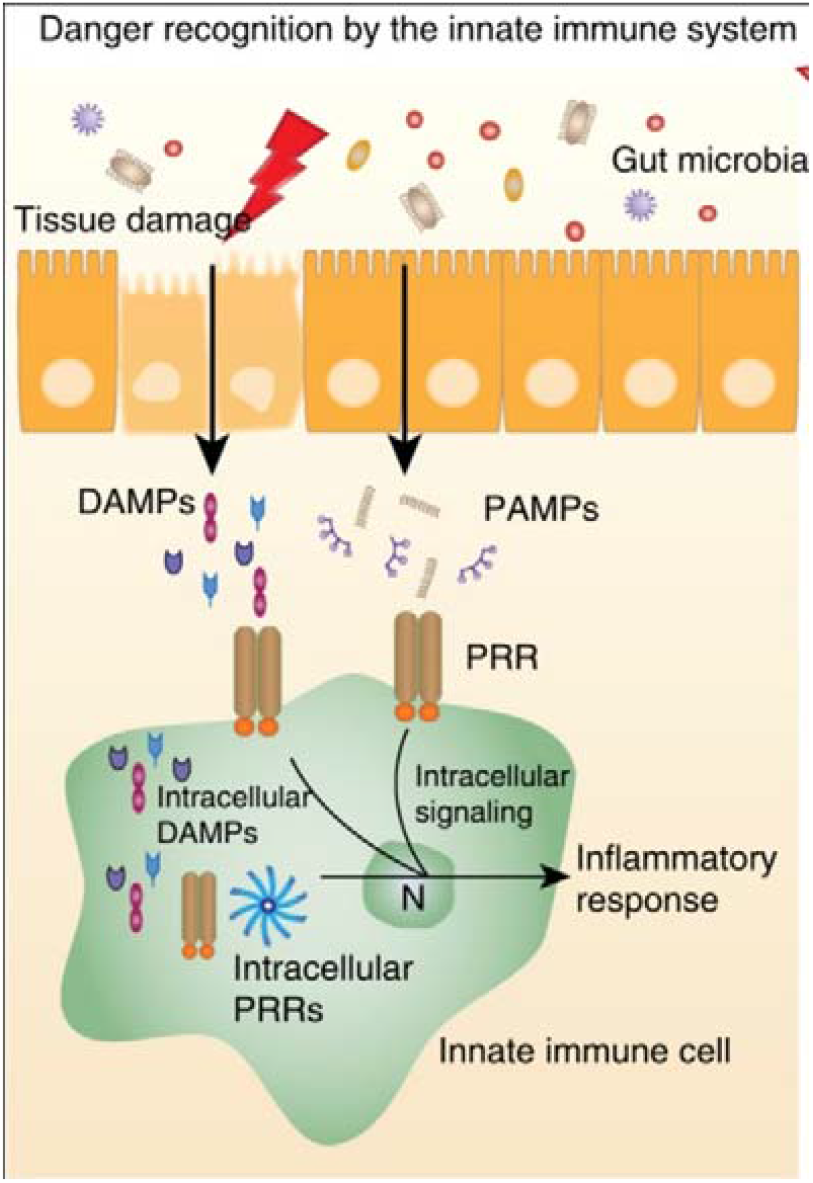

a. **How are DAMPs released?** Under physiological conditions, DAMPs reside intracellularly or are sequestered in the extracellular matrix and are thus hidden from recognition by innate immune cells bearing PRRs. In response to perceived danger such as tissue damage, DAMPs are liberated extracellularly, serving to signal danger to the host and promoting inflammation and repair processes. These processes are initially beneficial and protective, but in prolonged or significant DAMP release, they can be harmful.
b. **The relative importance of the specific DAMPs** DAMPs encompass a diverse range of biomolecules. Some are highly pro-inflammatory such as mitochondrial DNA (mtDNA), mitochondrial formylated peptides (mtFP) whereas others such as s100a8/9 (also called calprotectin) have less clear roles.
c. **What inflammatory pathways they activate?** Specific DAMPs have distinct roles on established inflammatory pathways. Our recent work has focused on mitochondrial DAMPs on toll-like receptor (TLR)-9 and Formylated Peptide Receptor (FPR)-1 signalling

### 1.4 What are mitochondrial DAMPs?

Recently, we found that DAMPs arising from the mitochondria are increased in patients with active IBD. Mitochondria are the ‘batteries’ or ‘powerstations’ that reside within and provide energy for living cells. They have evolved from bacteria around 2-3 billion years ago. As such, the mitochondria have many similarities with bacteria. When our immune cells encounter mitochondria that are released, they confuse them with bacteria, become activated and trigger a prolonged inflammatory response, which is destructive to our own tissue. Of interest, we showed that these signals are mitochondrial DNA and fragments of their protein, called formylated peptides.

Mitochondrial DAMPs can also be released in severe acute tissue damage or inflammation (for example, in major trauma or sepsis respectively). Their roles as biomarkers in chronic immune-mediated conditions such as IBD have not been fully evaluated.

#### 1.4.1 Mitochondrial DAMPs in IBD

We recently found two classes of mitochondrial DAMPs: DNA and formylated peptides that were significantly higher in IBD (prospective cohort of 67 UC and 30 CD patients)^14^. In CD, Plasma mtDNA levels were significantly higher in CD (136.7 copies/µl [IQR 88.0-370.9]) compared to non-IBD controls (61.5 copies/µl [IQR 32.8-104]) (p<0.0001). Higher mtDNA levels were observed in those with severely active CD (159.1 copies/µl [IQR 90.17-421]) compared to those in clinical remission (79.92 copies/µl [IQR 30.94 – 145.9] (p=0.04). In our overall IBD cohort, we found that mtDNA levels were significantly correlated with severe disease markers C-reactive protein (r=0.33, p<0.0001), albumin (r=-0.32, p<0.0001), and white cell count (*r*=0.37, p<0.0001). We also detected significantly higher levels of stool mtDNA and mitochondrial formylated peptides (that activates the pro-inflammatory formylated peptide receptor-1, [FPR1]) during active disease (vs. remission).

There are now approximately 260 genes that are associated with increased susceptibility to IBD in humans. Many of these IBD genes have direct roles in regulating mitochondrial function^15,17^. One of the strongest genetic signals from GWA-studies arises from genes regulating autophagy (*Atg16l1, Irgm, Atg5, Lrrk2, Park7*)^15,17^. Autophagy is the major biological process to remove damaged mitochondria. We (and others) have shown that mice models with autophagy defects (*Irgm*^*-/-*^, *Atg16l1*^*-/-*^, *Xbp1*^*-/-*^, *NLRP6*^*-/-*^ *and mdr1a*^*-/-*^) accumulate damaged mitochondria in the gut^18-21^. Failure to clear increases the burden and inflammatory potential of these damaged mitochondria. Related studies show that this results in higher pro-inflammatory mtDAMP release^22,23^. This pathologic process is amplified in defective autophagy. Hence, genetic susceptibility may influence the level of mitochondrial DAMPs release, which in turn, play a role in determining the severity of gut inflammation. Mitochondrial DAMPs therefore, also identifies the pathologic process IBD.

## 2 WHY IS THE MUSIC STUDY NEEDED?

We hypothesise that mitochondrial DAMPs are good mechanistic biomarkers for mucosal inflammation and healing in IBD.

Complete mucosal healing (total resolution and absence of ulcerations in the gut) is the most sought-after treatment target with the best long-term implication in prognosis.

Up to now IBD clinicians rely on (1) clinical symptoms (how they feel, their bowel habit, presence of blood in stools), (2) clinical tests such as stool calprotectin (FC) and blood C-reactive protein (CRP) to inform both themselves and the patients, how well the drug treatment is working and importantly, whether the ulcers and inflammation seen in the gut lining have healed or not.

Current evidence shows that these approaches are not fully informative. For example, 30% of patients with significant subjective improvement in their symptoms following treatment of active CD have evidence of active inflammation in their gut lining when further assessed with an ileo-colonoscopy. Blood and stool tests to predict mucosal healing are only useful in around 60-70% and very limited, in guiding the doctors in how severely inflamed the bowel wall is during active IBD.

Direct visualisation using ileo-colonoscopy or flexible sigmoidoscopy is the most accurate approach to assess disease activity and mucosal healing in response to medical treatment. By knowing precisely, how the gut wall inflammation is responding to treatment, the clinician can accurately manage the IBD patient (by either changing the dose and type of treatment, and whether to carry on with expensive, strong medications with potentially serious side effects). However, in the real world, follow-up endoscopic tests are difficult to carry out as they are expensive and we lack the capacity to undertake these examinations within NHS.

## 3 PROJECT GOALS

The main goal for the MUSIC study is to investigate the role of mitochondrial DAMPs in the clinic as an indicator of gut inflammation and subsequent mucosal healing in response to medical treatment in IBD.

Secondly, we will carry out further scientific studies using blood, stool, saliva and gut biopsy samples to investigate how mitochondrial DAMPs (and all known biomarkers and biological data such as genetics) contribute to the abnormal development of gut inflammation in IBD.

### 3.1 Primary research questions

- Do mitochondrial DAMPs predict the activity and severity of IBD-inflammation?
- Does normalisation of mitochondrial DAMPs reflect complete endoscopic mucosal healing in IBD?
- How do mitochondrial DAMPs compare to current biomarkers (FC, CRP) and clinical symptoms (HBI/Mayo Score) in assessing IBD inflammation and mucosal healing?
- Can we develop a simple decision-making model to predict mucosal healing based on mitochondrial DAMPs, together with relevant biological data such as genetics, blood transcriptomics, microbiome; and current clinical biomarkers such as calprotectin, faecal haemoglobin and blood CRP?

### 3.2 Secondary research questions

- How are mitochondrial DAMPs released from cells in the IBD gut?
- What types of cells are important in mitochondrial DAMP release? They are many forms of inflammatory cells in affected IBD gut (e.g. macrophage, epithelial, neutrophils). We think different cell types may contain more inflammatory DAMPs.
- Which type of mitochondrial DAMPs are important in causing inflammation? Can mitochondrial DAMPs pinpoint a specific underlying genetic susceptibility (e.g. autophagy) or inflammatory mechanism in IBD?

### 3.3 Rationale

Our focus is to investigate mitochondrial DAMPs’ utility in two clinically relevant scenarios: (a) How severe or active is the disease? (b) How well are we treating IBD? - with endoscopic endpoints of mucosal inflammation and healing respectively. We will use the Simple Endoscopic Score for Crohn’s Disease (SES-CD) and Endoscopic Mayo Score (eMS) for CD and UC respectively. Both have been validated and used widely in research and in clinical trials. By using these objective endoscopic endpoints, we can test mtDAMPs (and in combination with current biomarkers FC and CRP) across a range of mucosal inflammation (full healing to severe).

In addition to this, we will investigate if mitochondrial DAMPs can identify a subclinical pathogenic mechanism (e.g. [a] defective autophagy to clear damaged mitochondria; [b] de-regulated innate immune response to mitochondrial DAMPs.) These data will pave the way for future use of mitochondrial DAMP biomarkers as part of a stratified approach for new treatments targeted at mitochondrial DAMPs and their downstream inflammatory mechanisms in IBD.

## 4 STUDY POPULATION

Presently within usual NHS care, all patients with active IBD, especially those to be initiated on biologic or immunomodulator treatment, are followed up where they will have documentation of disease activity (Harvey Bradshaw Index [HBI] or UC Mayo Score [MS], stool calprotectin, C-reactive protein, albumin and blood count) to assess their well-being and response to medical treatment.

With MUSIC, our patients will be followed up prospectively (aligning the usual NHS clinical care above) and will receive additional endoscopic follow-up to assess mucosal healing in response to medical treatment. Thus, the MUSIC study will incorporate a prospective endoscopic evaluation of mucosal inflammation and mucosal healing into IBD clinics.

### Protocol

All participants will have active IBD at the time of recruitment.

We aim to capture a wide spectrum of IBD patients with active disease, hence strict criteria for IBD disease severity/extent/activity ***is not applied***.

However, suitable potential participants must have active IBD based on clinical evaluation of referring clinician and any one of the below from investigations which have been carried out within 6 weeks of screening:

1. FC level of >100ug/g
2. Blood CRP >5mg/l
3. Endoscopic, radiological or histological evidence of active IBD

There will be 2 main groups:

1. IBD patients with acute flare requiring medical treatment (corticosteroids, immunomodulator or biologics)
2. IBD patients on established medical treatments (biologics or immunomodulators) that require a change due to poor control of disease activity

## 5 PATIENT SELECTION AND ENROLMENT

### Patient screening and recruitment

Potential participants will be identified by responsible NHS IBD clinicians and then be referred to the MUSIC research team or local Principal Investigator in typical scenarios below:

1. IBD patients referred for the initiation of OR a switch in biologic treatment or immunomodulator treatment due to active disease
2. IBD patients who are newly diagnosed (following ileo-colonoscopy or flexible sigmoidoscopy, either as outpatient or inpatient) requiring corticosteroids (oral or intravenous), biologic treatment or immunomodulator treatment
3. IBD patients who are on medical follow-up, requiring endoscopic re-assessment of IBD activity due to worsening of their IBD symptoms
4. IBD patients who have contacted IBD service with symptoms suggestive of an IBD flare.

Following referral, the local MUSIC research teams will screen the suitability of the patient and send the potential participant a patient invitation letter and patient information sheets by post or email or give this to them in person.

The participant will have at least 24 hours to read this information and decide whether to take part. For in-patients, participants will have at least 6 hours to decide.

The first approach to the participant will be performed by the MUSIC research team (gastroenterologist/clinical research fellow or nurse) following:

1. IBD clinical review (in physical or virtual clinics; OR IBD Helpline contact by patients) OR during in-patient stay for management of active IBD
2. IBD patients’ hospital appointments for counselling for biologic therapy

Following initial screening and PIS allocation, our MUSIC research team will engage with suitable subjects in a dedicated research clinic. Prior to invitation, the MUSIC research team will contact potential participants by telephone or email to arrange a suitable time for clinic appointment.

If the participant agrees, the consenting process will take place during the appointment, and if the patient is willing to participate, sign the consent form. Following this, the MUSIC research team will plan subsequent steps for MUSIC study entailed below (Figure 1).

At this point (baseline), participants will be asked to fill a simple questionnaire, receive guidance to report on their clinical symptoms and have their index blood, saliva and stool tests taken.

Appointments for follow-up ileo-colonoscopy or flexible sigmoidoscopy will be arranged.

Follow-up visits at 3, 6, 9 and 12 months can be conducted either in a dedicated research, telephone or virtual clinic.

Appointments for further blood and stool tests will be arranged for participants at dedicated clinic appointments or home visits at a convenient time for the participant.

Follow-up clinical information on patient reported outcomes of well-being and IBD activity will be collected in clinic visits, by post or by a secured questionnaire that is linked to the study database.

No patient identifiable information is carried on the questionnaire. Details from the questionnaire will be transcribed where necessary and stored on an established database held on a University of Edinburgh server. A Caldicott guardian permission to hold information on this database from NHS Lothian records will be sought.

A study specific website will be created to keep participants informed of study news. Social media will also be used to communicate study information online.

### 5.1 Number of participants

We aim to recruit 250 IBD participants from participating gastroenterology units in Scotland.

### 5.2 Inclusion Criteria

1. All patients must be able to give consent and aged 16 years old and over.
2. All patients must have a diagnosis of IBD (CD or UC)
3. All patients must have active IBD at the time of screening:
  - Active IBD symptoms by referring clinician’s judgement in addition to one of the below criteria (within 6 weeks of screening):
    - FC level of >100ug/g
    - Blood CRP >5mg/l
    - Endoscopic, radiological or histological evidence of active IBD
4. All IBD patients with disease involvement that is amenable for endoscopic assessment of mucosal healing. This includes:
  - CD patients with previous ileal or colonic surgical resection
  - CD patients with perianal disease where ileo-colonoscopy or sigmoidoscopy are not contraindicated
  - CD patients with ileal involvement only where endoscopic disease activity can be recorded
5. All IBD patients will require a recent ileo-colonoscopy or flexible sigmoidoscopy within 6 weeks of recruitment that has:
  - Clear documentation of endoscopic disease activity and extent (SES-CD and Rutgeert’s score for CD; Mayo Score or UCEIS for UC)
  - Photographs of endoscopic mucosal IBD disease activity
  - If there is not a recent ileo-colonoscopy or flexible sigmoidoscopy, the participant will be asked to undergo an ileo-colonoscopy or flexible sigmoidoscopy at baseline.
6. If patients have undergone an ileo-colonoscopy or flexible sigmoidoscopy within 6 weeks but with an endoscopic report that is insufficient in endoscopic disease activity data as per (5), potential participant can still be considered providing there is:
  - Supporting objective evidence of IBD disease activity (FC, CRP) **within 2 weeks** of index ileo-colonoscopy or flexible sigmoidoscopy.

### 5.3 Exclusion Criteria

1. IBD patients with severe/fulminant disease at screening:
  - Subjects with colitis fulfilling the Truelove and Witts’ criteria (stool frequency >6/24 hours with **all** of the features of fever >38C, pulse rate >100 beats per minute, blood haemoglobin <105 g/l, albumin <30g/l)
  - Subjects displaying evidence of toxic megacolon (transverse colon diameter >6m on plain abdominal X-ray with accompanying full radiological report). Note – abdominal X-ray will be carried out if clinically indicated by referring clinician
  - Evidence of significant bowel obstruction, abdominal sepsis, abscess formation and fistula formation (bowel or perianal) as documented by referring clinician that is supported by clinical, radiological and blood laboratory investigations
2. Referring clinician’s judgement where surgical intervention (colectomy or resection) is deemed likely within 3 months of screening
3. Evidence of intestinal dyplasia or malignancy (histologic, endoscopic or radiologic)
4. UC patients with limited involvement of the rectum (<15cm – proctitis)
5. UC patients who have had a colectomy (total and subtotal)
6. UC patients with an ileo-anal pouch
7. IBD (UC, CD or IBD-U) with an intestinal stoma
8. Patients where ileo-colonoscopy or flexible sigmoidoscopy are contra-indicated (e.g. significant co-morbidities e.g. cardiovascular, respiratory, cancer, renal failure; and pregnancy)
9. Participants where there are limitations to language communication where there is a potential issue where information sheet cannot be reliably understood and/or the subject cannot provide full informed consent.

### 5.4 Co-enrolment

Co-enrolment to other research studies, including drug, interventional and long-term follow-up studies will be permitted if this has been agreed and documented by the Chief Investigators of co-enrolling studies.

Any co-enrolment will follow the Sponsor Guidelines GL001.

### 5.5 Study groups

The recruitment and subsequent clinical data and sample collection will take place at 0, 3, 6, 9 and 12 months.

If a study participant is admitted to hospital for further in-patient management during the study period, our MUSIC research team will record the clinical and laboratory investigations pertaining to the hospital admission event. Further research stool and blood tests will also be carried out during the hospital admission.

At 0, 3, 6, 9 and 12 months, these participants will provide clinical activity score, FC, CRP; and as part of MUSIC study, provide additional blood and stool for biomarker measurements.

At 3-month (±1 months) [*and at 12 months (at local PI’s discretion)*] following baseline, all participants will undergo a further ileo-colonoscopy or flexible sigmoidoscopy (for UC).

Participants may have had a recent index ileo-colonoscopy or flexible sigmoidoscopy as part of their routine clinical care prior to recruitment (<6 weeks before recruitment).

If not, they will have one carried out by our research team as part of the MUSIC study to assess the extent and severity of mucosal inflammation at baseline.

### 5.6 Study investigations

Some study assessment results may be obtained from participants’ medical records.

**Table 1.**
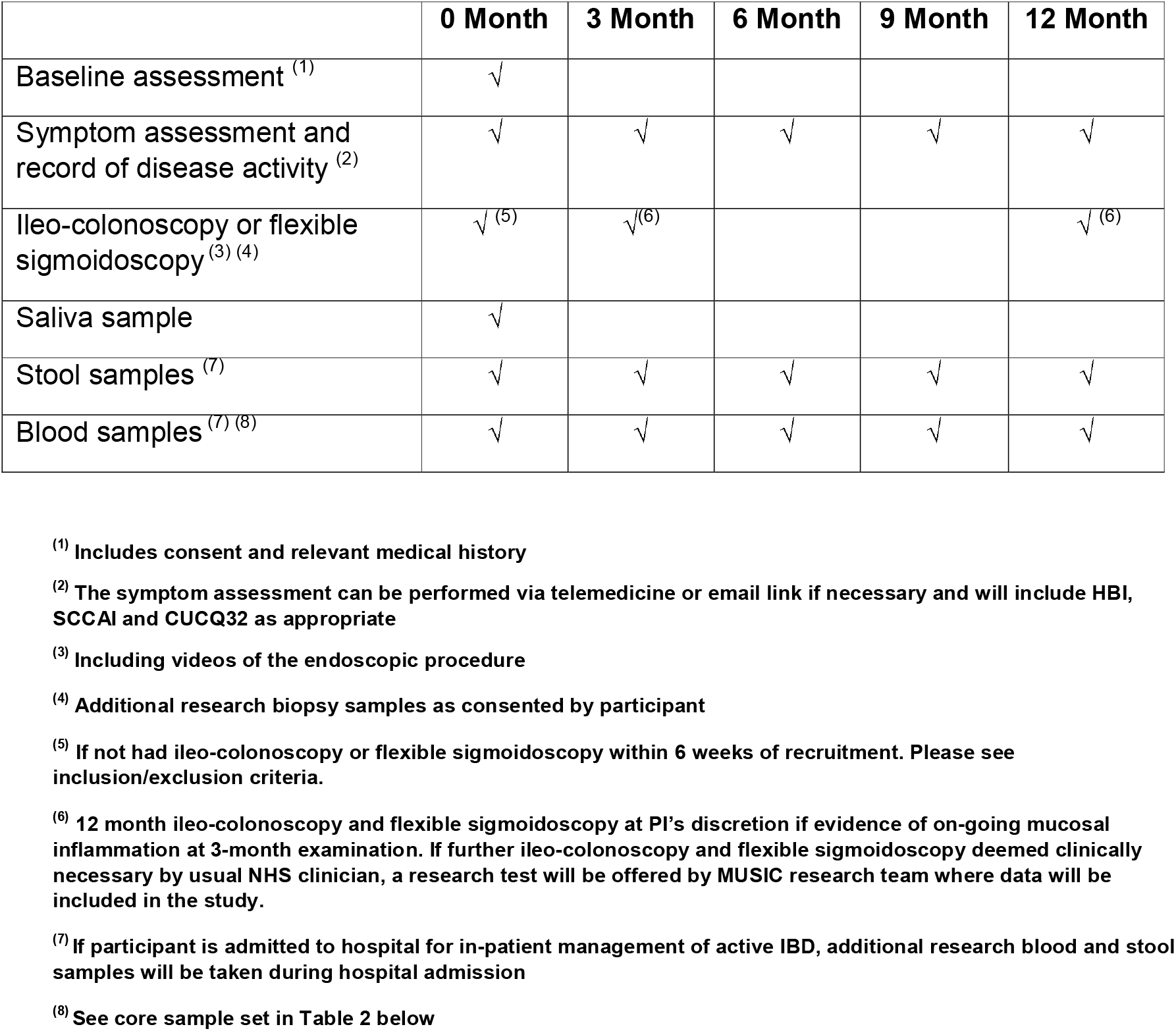

Participants may be contacted by the MUSIC research team by email, phone or text to remind them of appointments or sample collection.

#### 5.6.1 Study Samples

Samples will be linked to the MUSIC system by use of patient-specific barcodes or QR codes generated by the study team.

**Table 2.**
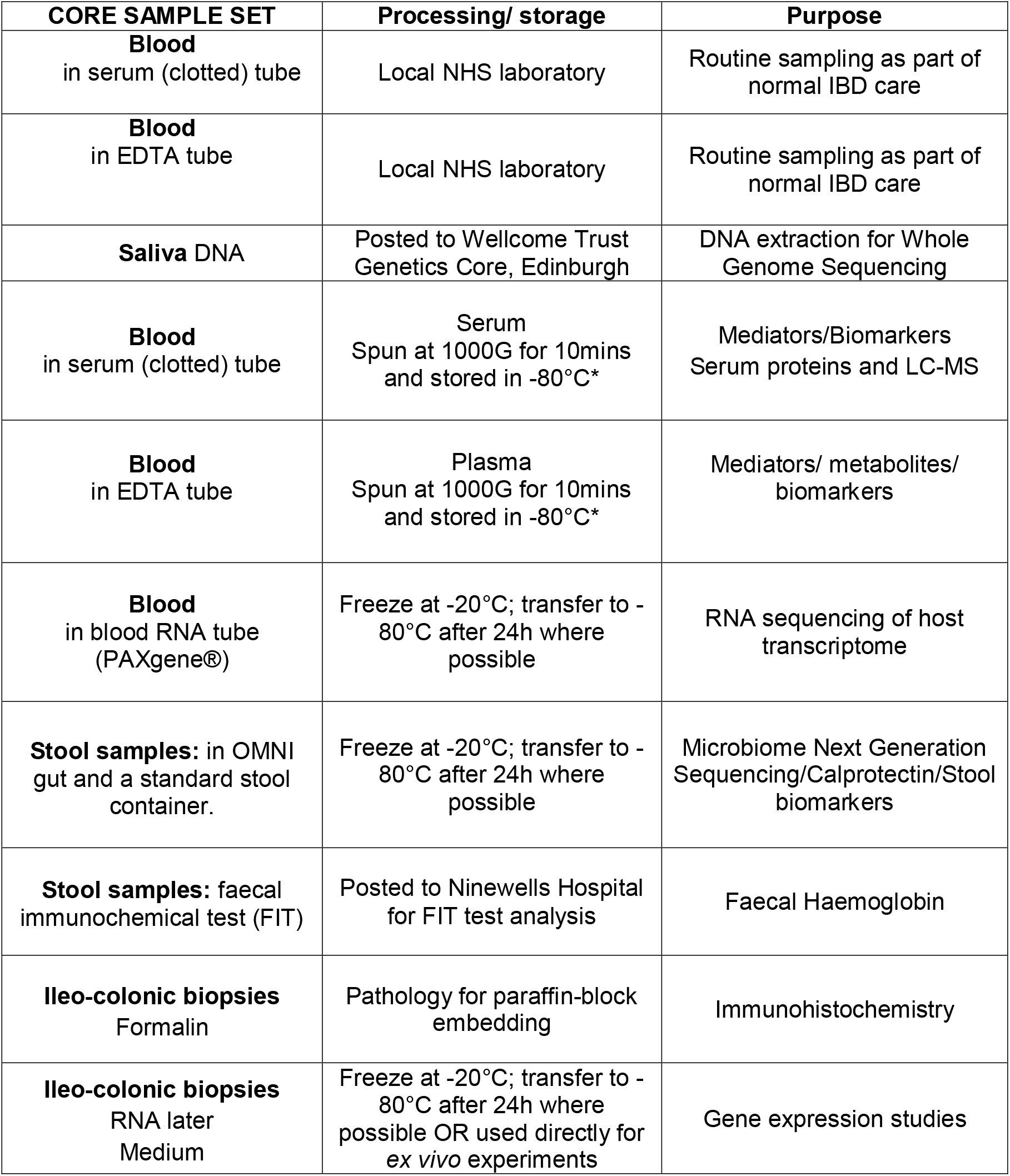

| CORE SAMPLE SET | Processing/ storage | Purpose |
| --- | --- | --- |
| **Blood**<br>in serum (clotted) tube | Local NHS laboratory | Routine sampling as part of normal IBD care |
| **Blood**<br>in EDTA tube | Local NHS laboratory | Routine sampling as part of normal IBD care |
| **Saliva DNA** | Posted to Wellcome Trust Genetics Core, Edinburgh | DNA extraction for Whole Genome Sequencing |
| **Blood**<br>in serum (clotted) tube | Serum<br>Spun at 1000G for 10mins and stored in -80°C* | Mediators/Biomarkers<br>Serum proteins and LC-MS |
| **Blood**<br>in EDTA tube | Plasma<br>Spun at 1000G for 10mins and stored in -80°C* | Mediators/ metabolites/ biomarkers |
| **Blood**<br>in blood RNA tube (PAXgene®) | Freeze at -20°C; transfer to -80°C after 24h where possible | RNA sequencing of host transcriptome |
| **Stool samples:** in OMNI gut and a standard stool container. | Freeze at -20°C; transfer to -80°C after 24h where possible | Microbiome Next Generation Sequencing/Calprotectin/Stool biomarkers |
| **Stool samples:** faecal immunochemical test (FIT) | Posted to Ninewells Hospital for FIT test analysis | Faecal Haemoglobin |
| **Ileo-colonic biopsies**<br>Formalin | Pathology for paraffin-block embedding | Immunohistochemistry |
| **Ileo-colonic biopsies**<br>RNA later<br>Medium | Freeze at -20°C; transfer to -80°C after 24h where possible OR used directly for *ex vivo* experiments | Gene expression studies |

**SALIVA:** During study period, participant will be asked to provide 1 saliva sample, which will be collected for DNA sampling.**BLOOD SAMPLES:** At each time point above, ∼40mls of blood (approximately 4-5 standard blood tubes) will be taken either at usual IBD, research or phlebotomy clinic during the hospital visit or within the community.

##### ILEO-COLONOSCOPY (or FLEXIBLE SIGMOIDOSCOPY)

A. We will perform an endoscopic examination of the bowel lining in all participants. Two types of approaches are available: ileo-colonoscopy where the entire large bowel and lower portion of the small bowel (ileum) are examined OR flexible sigmoidoscopy where only the left side of the large bowel is examined. Flexible sigmoidoscopy is a shorter test in duration.
B. In all participants, we will perform an ileo-colonoscopy EXCEPT in participants with UC where a baseline test has shown that the entire extent of the large bowel inflammation can be adequately examined by a flexible sigmoidoscopy.
C. If a participant requires an ileo-colonoscopy or flexible sigmoidoscopy at baseline, our study will carry out this procedure as part of our research study (as baseline).
D. If a participant has had an ileo-colonoscopy as part of routine clinical care within 6 weeks of study approach, he/she can still be recruited into the study, providing that the ileo-colonoscopy has adequate record of endoscopic activity of mucosal inflammation (SES-CD/Mayo Score) with supporting image data. We will use this ileo-colonoscopy as baseline (0 month).
E. At 3 (±1) month time point, each participant will undergo a follow-up ileo-colonoscopy or flexible sigmoidoscopy to record endoscopic activity of mucosal inflammation (SES-CD/Mayo Score).
F. If at 3-month time-point, ileo-colonoscopy or flexible sigmoidoscopy test shows on-going evidence of mucosal inflammation (i.e. no complete mucosal healing or improvement from index baseline test), a further examination will be arranged for 12-month (+/- 1 month) time point.
G. In certain clinical situations, if NHS clinical team require a further ileo-colonoscopy to assess IBD activity, our study will carry out this procedure as part of our research study.

**Hence, all participants will have at least two endoscopic tests (ileo-colonoscopy or flexible sigmoidoscopy) during a 12-month follow-up period**. Participants will be posted a bowel preparation agent by post by local GI clinical teams, fasted overnight and may receive intravenous sedation as part of the procedure.

##### MUSIC STUDY

Ileo-colonoscopy/Flexible sigmoidoscopy follow-up plan

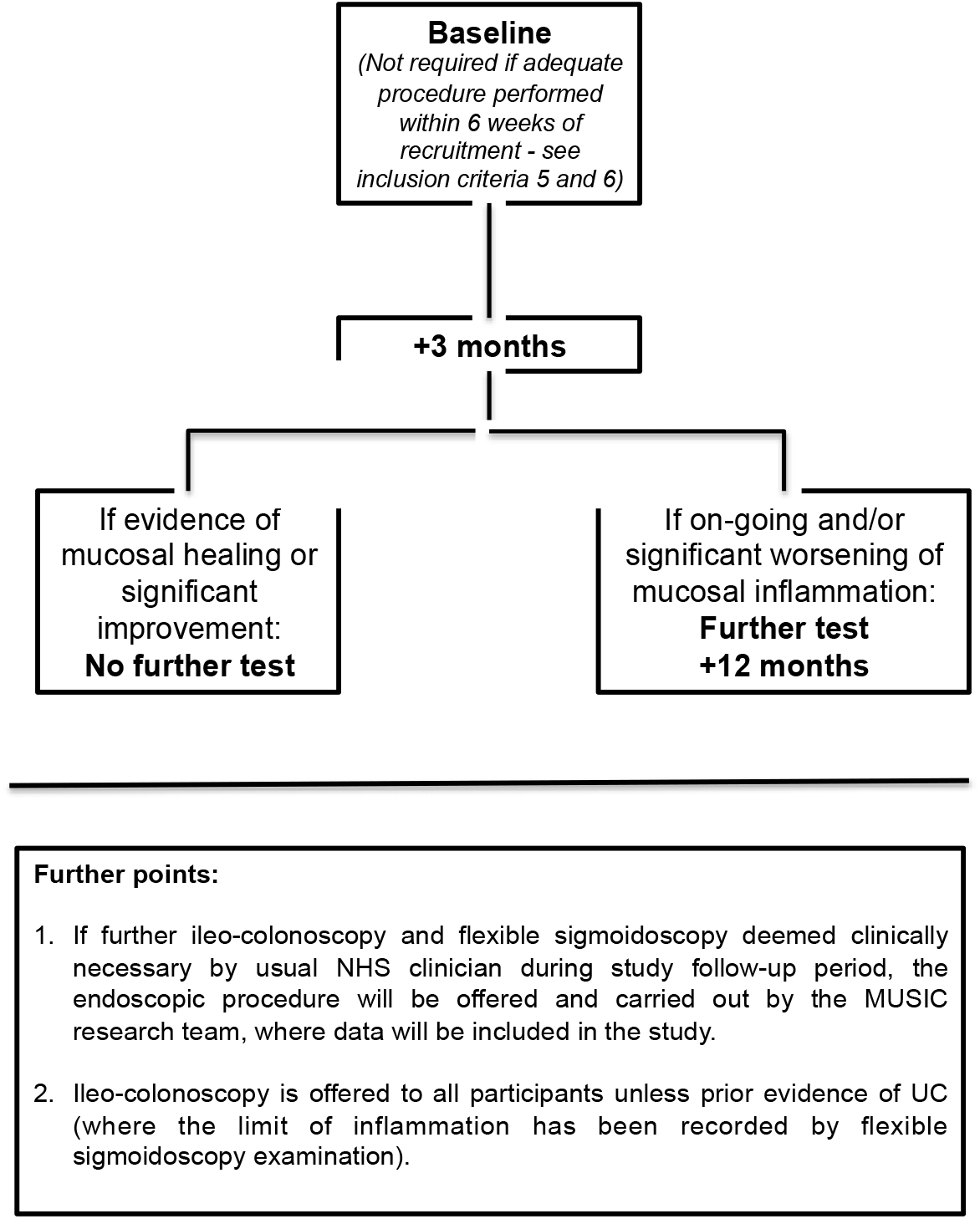

##### ILEO-COLONOSCOPY (or FLEXIBLE SIGMOIDOSCOPY) SAMPLES

For research, up to 16 (usually 8-10) pinch biopsies will be collected. Standard research sampling involves: 4 biopsies (2 for formalin fixation and 2 for RNA-later) from ileum (the small bowel), caecum, transverse colon and rectum respectively. Total of 16 biopsies.

These research biopsies will be in addition to routine biopsies taken for histopathological evaluation of IBD: Up to 16 pinch biopsies from ileum and across the colonic regions.

As there are extra research biopsies taken, there is a slightly higher risk of excessive bleeding (<1%). Most bleeding will settle spontaneously and if there is more bleeding than expected, this can usually be treated by cauterisation or clipping during the same procedure by the Endoscopist.

##### ILEO-COLONOSCOPY (or FLEXIBLE SIGMOIDOSCOPY) IMAGES

Photographic and video images of the inflamed and uninflamed bowels will be taken during ileo-colonoscopy for further analysis to provide an objective score for disease activity and severity. Photographic images are usually taken during standard ileo-colonoscopy.

#### 5.6.2 In selected patients

##### BLOOD SAMPLES

In approximately 40 patients, more blood is necessary to carry out studies where we need to isolate blood immune cells to study the effect of DAMP release and inflammatory actions. Here, we expect to study 10-20 participants with anticipated high DAMP release (IBD patients with more active disease – frequent diarrhoea, high inflammatory blood markers). In parallel, there is a requirement for ∼10-20 IBD patients with inactive, quiescent disease (no diarrhoea, normal routine blood markers). Hence, in total ∼30-40 participants. In this setting, a maximum of 100mls of blood is required. When we are required to take the large sample of blood, we will make this clear at the time of recruitment and re-emphasise that this part is optional

##### SURGICAL SAMPLES

If during duration of MUSIC study follow-up, surgery to remove affected part of IBD bowel is necessary, surplus tissue may be obtained from specimens of the large bowel (colon) and lower small bowel (ileum) that have been surgically removed either at the time of operation or during histopathological evaluation.

### 5.7 Withdrawal of study participants

Participants are free to withdraw from the study at any point or the Investigator can withdraw a participant. If withdrawal occurs, the primary reason for withdrawal will be documented in the participant’s case report form, if possible.

### 5.8 Storage and analysis of samples

All research samples will be anonymised and given a study code. Samples will be transferred between participating sites to allow for storage or analysis by members of the research team.

**SALIVARY DNA** will be posted directly to the Gut Research Unit, University of Edinburgh for processing and storage.

**RESEARCH STOOL SAMPLES** will be posted directly to Edinburgh (OmniGUT) and Dundee (qFIT) for dedicated analysis in special envelopes.

**RESEARCH BLOOD SAMPLES** will be processed and stored initially at the participating centres using an agreed Standard Operating Procedure (SOP).

**ENDOSCOPIC BIOPSIES** in RNAlater are processed and stored in appropriate temperature controlled storage using an agreed Standard Operating Procedure (SOP).

**ENDOSCOPIC BIOPSIES** are also stored in formalin and sent to local Pathology Department to be processed initially. The coded pathology blocks will then be sent to and stored at the Gut Research Unit, Centre for Inflammation Research, Queen’s Medical Research Institute, University of Edinburgh At 3-4 monthly intervals, depending on progress of recruitment, these samples will then be processed as a batch to process supernatants (**STOOL**), plasma and serum (**BLOOD**) and RNA/protein extraction (**ENDOSCOPIC BIOPSIES**) for planned experiments.

## 6 DATA COLLECTION AND MANAGEMENT

Research data will be collected using the MUSIC application system designed and developed in the REDCap platform, which is maintained by the University of Edinburgh IT department based at the Institute of Genetics and Molecular Medicine (IGMM) at Western General Hospital campus in Edinburgh.

In the MUSIC system, we will use the REDCap secure web application for managing non patient identifiable information and questionnaires. It is encrypted using AES-256 in counter mode and authenticating it using Poly1305-AES. Day to day management of this database is maintained by a dedicated Data Scientist in the MUSIC team and can only be accessed by members of the research team.

Hard copies of consent forms (with linked patient name and unique numbered code) and questionnaires (with allocated specific study ID) will be filed separately in locked filing cabinets in secure locations at each research site.

Routine clinical data on IBD activity during the follow-up period will be updated in the MUSIC database.

Mobile data collection devices will be used by clinical and laboratory staff to complete the data entry on the MUSIC system. In these cases, the relevant pages accessed using these mobile devices will not contain or be linked to any patient identifiable information.

All biological samples collected will be kept within the Gut Research Unit, Centre for Inflammation Research, Queens Medical Research Institute, University of Edinburgh and Wellcome Trust Clinical Research Facility, Western General Hospital under the oversight of the Lothian Gastroenterology Bioresource, University of Edinburgh and be processed every 2-3 months; or for certain experimental work, be used immediately in the Gut Research Laboratories, Centre for Inflammation Research, University of Edinburgh. Here, we will carry out new biomarker analysis from blood, stools and biopsies working together and in combination with our studies in Edinburgh. Samples and related clinical data may be shared with other researchers for use in other related projects on agreement with the Chief Investigator. At the end of the study, anonymised samples that are not directly used by our research will be transferred to South East Scotland SAHSC BioResource under the guardianship of NHS Lothian or disposed in accordance to the Human Tissue Authority Code of Practice.

All ileo-colonoscopic images/videos will be anonymised and coded according to allocated specific Study ID. All images/videos will be stored in a secure database in the University of Edinburgh server.

### 6.1 Personal Data

The following personal data will be collected as part of the research:

CHI number, date of birth, initials, gender, age and family history of relevant medical conditions will be collected. Patient phone numbers and email addresses may also be collected if they wish to complete their study questionnaires via email link or be reminded of samples or appointments by text message.

All data is stored on an established encrypted database held as above. The electronic database has the recommended encryption (AES256) and can only be accessed by members of the research team. The key or code that links study ID no to CHI or identifiable personal data will be stored in a separate encrypted database also held on a University of Edinburgh server. Only the Chief Investigator (Dr Gwo-tzer HO) and named PIs will have the code to for this database.

Hard copies of consent forms, data collection forms and questionnaires (with allocated specific study ID) will be filed separately and securely in locked filing cabinets at each study site.

### 6.2 Transfer of Data

Data from this project may be provided to researchers running other research studies in this organisation and in other organisations. These organisations may be universities, NHS organisations or companies involved in health and care research in this country or abroad. Information will only be used by organisations and researchers to conduct research in accordance with the <u>UK Policy Framework for Health and Social Care Research</u>. This data could be used for research in any aspect of health or care, and could be combined with information from other sources held by researchers, the NHS or government.

### 6.3 Data Custodian

University of Edinburgh and NHS Lothian are the co-sponsors and will act as data custodians for this study based in Scotland. We will use information from participants and/or their medical records in order to undertake this study and the co-sponsors will act as the data controller for this study. This means that University of Edinburgh and NHS Lothian are responsible for looking after information and using it properly. The co-sponsors will keep identifiable information about participants for 5 years.

The data custodian is the person who will be responsible for the use, security and management of all data generated by the study.

### 6.4 Data Controller

The data controller is University of Edinburgh

### 6.5 Data Breaches

The online database is hosted on a secure University of Edinburgh server. Any data breaches will be reported to the University of Edinburgh who will onward report to the relevant authority.

### 6.6 Identifiable Data for future research

University of Edinburgh and NHS Lothian are the co-sponsors for this study based in Scotland. The co-sponsors are responsible for any identifiable information about participants for 5 years and are strictly governed by <u>UK Policy Framework for Health and Social Care Research</u>.

## 7 ADVERSE EVENTS

Adverse events (AE) and serious adverse events (SAE) are defined in ACCORD SOP CR006. The current version of this SOP can be downloaded from the ACCORD website. http://accord.scot/research-access/resources-researchers/ We propose to report centrally only those SAEs which directly result from the conduct of the study (e.g. venepuncture for research purposes). Any AE or SAE which is likely to have or even possibly arisen from the conduct of the study, but not those due to natural disease progression will be handled according to SOP CR006.

## 8 OVERSIGHT ARRANGEMENTS

### 8.1 INSPECTION OF RECORDS

Investigators and institutions involved in the study will permit trial related monitoring and audits on behalf of the sponsor, REC review, and regulatory inspection(s). In the event of audit or monitoring, the Investigator agrees to allow the representatives of the sponsor direct access to all study records and source documentation. In the event of regulatory inspection, the Investigator agrees to allow inspectors direct access to all study records and source documentation.

### 8.2 STUDY MONITORING AND AUDIT

The ACCORD Sponsor Representative will assess the study to determine if an independent risk assessment is required. If required, the independent risk assessment will be carried out by the ACCORD Quality Assurance Group to determine if an audit should be performed before/during/after the study and, if so, at what frequency.

## 9 GOOD CLINICAL PRACTICE

### 9.1 ETHICAL CONDUCT

The study will be conducted in accordance with the principles of the International Conference on Harmonisation Tripartite Guideline for Good Clinical Practice (ICH GCP).

Before the study can commence, all required approvals will be obtained and any conditions of approvals will be met.

### 9.2 INVESTIGATOR RESPONSIBILITIES

The Investigator is responsible for the overall conduct of the study at the site and compliance with the protocol and any protocol amendments. In accordance with the principles of ICH GCP, the following areas listed in this section are also the responsibility of the Investigator. Responsibilities may be delegated to an appropriate member of study site staff.

#### 9.2.1 Informed Consent

The Investigator is responsible for ensuring informed consent is obtained before any protocol specific procedures are carried out. The decision of a participant to participate in clinical research is voluntary and should be based on a clear understanding of what is involved.

Participants must receive adequate oral and written information – appropriate Participant Information and Informed Consent Forms will be provided as documented earlier.

The oral explanation to the participant will be performed by the Investigator or qualified delegated person, and must cover all the elements specified in the Participant Information Sheet and Consent Form.

The participant must be given every opportunity to clarify any points they do not understand and, if necessary, ask for more information. The participant must be given sufficient time to consider the information provided. It should be emphasised that the participant may withdraw their consent to participate at any time without loss of benefits to which they otherwise would be entitled.

The participant will be informed and agree to their medical records being inspected by regulatory authorities and representatives of the sponsor(s).

The Investigator or delegated member of the trial team and the participant will sign and date the Informed Consent Form(s) to confirm that consent has been obtained. The participant will receive a copy of this document and a copy filed in the Investigator Site File (ISF) and participant’s medical notes (if applicable).

As highlighted in the earlier patient recruitment section, the process of consenting will be carried out by individuals within the Delegation Log for consent. This will include the Principal Investigator, named Investigators, MUSIC clinical research doctors and nurses.

In most circumstances, consenting process will be carried out within dedicated MUSIC outpatient clinic appointments that have been agreed and arranged (following referral). Otherwise, less commonly they will take place during dedicated clinic time following clinic appointments with routine consultant review or as an in-patient during concomitant hospital admission. Every effort will be made to minimize inconvenience for the participant.

#### 9.2.2 Study Site Staff

The Investigator must be familiar with the protocol and the study requirements. It is the Investigator’s responsibility to ensure that all staff assisting with the study are adequately informed about the protocol and their trial related duties.

#### 9.2.3 Data Recording

The Principal Investigator is responsible for the quality of the data recorded in the CRF at each Investigator Site.

#### 9.2.4 Investigator Documentation

The Principal Investigator will ensure that the required documentation is available in local Investigator Site files ISFs.

#### 9.2.5 GCP Training

For non-CTIMP (i.e. non-drug) studies all researchers are encouraged to undertake GCP training in order to understand the principles of GCP. However, this is not a mandatory requirement unless deemed so by the sponsor. GCP training status for all investigators should be indicated in their respective CVs.

#### 9.2.6 Confidentiality

All laboratory specimens, evaluation forms, reports, and other records must be identified in a manner designed to maintain participant confidentiality. All records must be kept in a secure storage area with limited access. Clinical information will not be released without the written permission of the participant. The Investigator and study site staff involved with this study may not disclose or use for any purpose other than performance of the study, any data, record, or other unpublished, confidential information disclosed to those individuals for the purpose of the study. Prior written agreement from the sponsor or its designee must be obtained for the disclosure of any said confidential information to other parties.

#### 9.2.7 Data Protection

All Investigators and study site staff involved with this study must comply with the requirements of the Data Protection Act 2018 with regard to the collection, storage, processing and disclosure of personal information and will uphold the Act’s core principles. Access to collated participant data will be restricted to individuals from the research team treating the participants, representatives of the sponsor(s) and representatives of regulatory authorities.

Computers used to collate the data will have limited access measures via user names and passwords.

Published results will not contain any personal data that could allow identification of individual participants.

## 10 STUDY CONDUCT RESPONSIBILITIES

### 10.1 PROTOCOL AMENDMENTS

Any changes in research activity, except those necessary to remove an apparent, immediate hazard to the participant in the case of an urgent safety measure, must be reviewed and approved by the Chief Investigator.

Amendments will be submitted to a sponsor representative for review and authorisation before being submitted in writing to the appropriate REC, and local R&D for approval prior to participants being enrolled into an amended protocol.

### 10.2 MANAGEMENT OF PROTOCOL NON COMPLIANCE

Prospective protocol deviations, i.e. protocol waivers, will not be approved by the sponsors and therefore will not be implemented, except where necessary to eliminate an immediate hazard to study participants. If this necessitates a subsequent protocol amendment, this should be submitted to the REC, and local R&D for review and approval if appropriate.

Protocol deviations will be recorded in a protocol deviation log and logs will be submitted to the sponsors every 3 months. Each protocol violation will be reported to the sponsor within 3 days of becoming aware of the violation. All protocol deviation logs and violation forms should be emailed to

Deviations and violations are non-compliance events discovered after the event has occurred. Deviation logs will be maintained for each site in multi-centre studies. An alternative frequency of deviation log submission to the sponsors may be agreed in writing with the sponsors.

### 10.3 SERIOUS BREACH REQUIREMENTS

A serious breach is a breach, which is likely to effect to a significant degree:

a. the safety or physical or mental integrity of the participants of the trial; or
b. the scientific value of the trial.

If a potential serious breach is identified by the Chief investigator, Principal Investigator or delegates, the co-sponsors must be notified within 24 hours. It is the responsibility of the co-sponsors to assess the impact of the breach on the scientific value of the trial, to determine whether the incident constitutes a serious breach and report to research ethics committees as necessary.

### 10.4 STUDY RECORD RETENTION

All study documentation will be kept for a minimum of 5 years from the protocol defined end of study point. When the minimum retention period has elapsed, study documentation will not be destroyed without permission from the sponsor. The sponsor SOP for study archiving will be followed.

### 10.5 END OF STUDY

The end of study is defined as the last participant’s last visit.

The Investigators or the co-sponsor(s) have the right at any time to terminate the study for clinical or administrative reasons.

The end of the study will be reported to the REC, and R+D Office(s) and co-sponsors within 90 days, or 15 days if the study is terminated prematurely. The Investigators will inform participants of the premature study closure and ensure that the appropriate follow up is arranged for all participants involved. End of study notification will be reported to the co-sponsors via email to

A summary report of the study will be provided to the REC within 1 year of the end of the study.

### 10.6 INSURANCE AND INDEMNITY

The co-sponsors are responsible for ensuring proper provision has been made for insurance or indemnity to cover their liability and the liability of the Chief Investigator and staff.

The following arrangements are in place to fulfil the co-sponsors’ responsibilities:

- The Protocol has been designed by the Chief Investigator and researchers employed by the University and collaborators. The University has insurance in place (which includes no-fault compensation) for negligent harm caused by poor protocol design by the Chief Investigator and researchers employed by the University.
- Sites participating in the study will be liable for clinical negligence and other negligent harm to individuals taking part in the study and covered by the duty of care owed to them by the sites concerned. The co-sponsors require individual sites participating in the study to arrange for their own insurance or indemnity in respect of these liabilities.
- Sites which are part of the United Kingdom’s National Health Service will have the benefit of NHS Indemnity.
- Sites out with the United Kingdom will be responsible for arranging their own indemnity or insurance for their participation in the study, as well as for compliance with local law applicable to their participation in the study.

## 11 REPORTING, PUBLICATIONS AND NOTIFICATION OF RESULTS

### 11.1 AUTHORSHIP POLICY

Ownership of the data arising from this study resides with the study team.

## Data Availability

All data produced in the present study are available upon reasonable request to the authors.

## Acknowledgement

This study is funded by The Leona M. and Harry B. Helmsley Charitable Trust (G-1911-03343)

## Ethical review

This study protocol was approved by the East of Scotland Research Ethics Service, Scotland, United Kingdom (Reference No. 19/ES/0087) on 17^th^ September 2019 and registered ClinicalTrials.gov as NCT04760964.

### LIST OF ABBREVIATION

ACCORD: Academic and Clinical Central Office for Research & Development - Joint office for The University of Edinburgh and Lothian Health Board
CHI: Community Health Index
CD: Crohn’s Disease
CDAI: Crohn’s Disease Activity Index
CI: Chief Investigator
CRF: Case Report Form
CRP: C-reactive protein
CTIMP: Clinical Trial of an Investigational Medicinal Product
CUCQ32: Crohn’s and Ulcerative Colitis Questionnaire
DAMPS: Damage Associated Molecular Patterns
DNA: Deoxyribonucleic Acid
ECTU: Edinburgh Clinical Trials Unit
EDTA: Ethylenediaminetetraacetic acid
eMS/MS: Endoscopic Mayo Score/Mayo Score
FBC: Full Blood Count
FC: Faecal (stool) calprotectin
FIT: Faecal Immunohistochemistry Test
FPR: Formylated Peptide Receptor
GCP: Good Clinical Practice
HBI: Harvey-Bradshaw Index
IBD: Inflammatory Bowel Disease
ICH: International Conference on Harmonisation
ISF: Investigator Site File
mtDNA: Mitochondrial DNA
mtFP: Mitochondrial Formylated Peptides
PI: Principal Investigator
PRR: Pattern Recognition Receptors
QA: Quality Assurance
RNA: Ribonucleic Acid
REC: Research Ethics Committee
SCCAI: Simple Clinical Colitis Activity Index
SES-CD: Simple Endoscopic Score for Crohn’s Disease
SOP: Standard Operating Procedure
TLR: Toll-like Receptor
TMF: Trial Master File
TNF: Tumour Necrosis Factor
U-E: Urea and Electrolytes
UC: Ulcerative Colitis

## Footnotes

1 Ileo-colonoscopy test is carried out to examine the entire large bowel (as colon) and final part of the small bowel (ileum). Approximately 100 and 10cm of the colon and ileum respectively can be inspected by ileo-colonoscopy. This is a necessary test for patients with CD as inflammation usually affects the ileum and anywhere along the large bowel. Flexible sigmoidoscopy test is carried out to examine the left side of colon (approximately 50cm of bowel lining can be visualized by this test) and is a shorter test in duration. As UC sometimes only affects the left side of the colon, flexible sigmoidoscopy is an adequate test to examine the extent and severity of UC.

